# Investigating the impact of weakly supervised data on text mining models of publication transparency: a case study on randomized controlled trials

**DOI:** 10.1101/2021.09.14.21263586

**Authors:** Linh Hoang, Lan Jiang, Halil Kilicoglu

## Abstract

Lack of large quantities of annotated data is a major barrier in developing effective text mining models of biomedical literature. In this study, we explored weak supervision strategies to improve the accuracy of text classification models developed for assessing methodological transparency of randomized controlled trial (RCT) publications. Specifically, we used Snorkel, a framework to programmatically build training sets, and UMLS-EDA, a data augmentation method that leverages a small number of existing examples to generate new training instances, for weak supervision and assessed their effect on a BioBERT-based text classification model proposed for the task in previous work. Performance improvements due to weak supervision were limited and were surpassed by gains from hyperparameter tuning. Our analysis suggests that refinements to the weak supervision strategies to better deal with multi-label case could be beneficial.

## Introduction

Incomplete reporting and lack of transparency are common problems in biomedical publications and may reduce the credibility of the findings of a study. These problems can have serious consequences, particularly in clinical research publications, since the evidence from these studies inform patient care and healthcare policy. In clinical research, randomized controlled trials (RCTs) are the most robust kind of primary research evidence regarding the effectiveness of therapeutic interventions^1^ and are a cornerstone of evidence-based medicine^2^. RCTs are time-consuming and expensive, and if inadequately designed, conducted, or reported, they lead to poor health outcomes and significant research waste^3^.

Reporting guidelines have been proposed to improve transparency and completeness of reporting for various types of biomedical studies. For example, the CONSORT statement focuses on RCT reporting^1, 4^, and consists of a 25-item checklist and a flow diagram. While endorsed by many high-impact medical journals, adherence to CONSORT remains inadequate^1, 5^ and difficult to enforce in practice, due to substantial workload it involves for journals. Manual CONSORT compliance checks before peer review have been shown to improve reporting quality^6^; however, they are difficult to scale and require significant domain expertise.

In previous work, we presented a corpus of 50 RCT publications manually annotated at the sentence level with fine-grained CONSORT checklist items and proposed a text mining approach to automate the task of transparency (reporting quality) assessment^7^. As a first step toward full transparency assessment, we developed sentence classification models to categorize sentences in the Methods sections of RCT publications into 17 methodology-related checklist items (e.g., Eligibility Criteria, Outcomes, Sequence Generation, Allocation Concealment). The best-performing model, based on BioBERT pretrained language model^8^, yielded reasonable performance on some items, particularly those that are commonly discussed in RCT Methods sections and thus are well-represented in the dataset. However, the results overall suffered from the relatively small size of the dataset and largely failed on the checklist items that are infrequently reported in RCT publications (e.g., Changes to Outcomes).

Annotated data is critical in training modern natural language processing and text mining (NLP) algorithms. In particular, deep neural network architectures heavily depend on large quantities of training data for learning model parameters. While recent pretrained language models, such as BERT^9^ and its variants, exhibit better sample efficiency and often work well even with relatively small datasets, the importance of annotated data has not diminished. High performance of BERT-based models in NLP tasks and the resulting standardization of architectures arguably underlines data scarcity as the primary bottleneck in NLP^10^. In response, weak supervision techniques have become increasingly popular, as they offer cheaper or more efficient ways to generate training data^10, 11^.

In this study, we investigated whether weak supervision techniques can be used to effectively label additional data and improve our sentence classification models for transparency assessment of RCT publications. More specifically, we focused on weak supervision using the Snorkel framework^10, 11^ and data augmentation based on the UMLS-EDA algorithm^12^ and used the labels that they generated as additional data for the BioBERT-based model reported in previous work^7^. The results show that weak supervision has limited effectiveness on our dataset, while at the same time indicating that hyperparameter tuning can have a more significant impact on model performance.

## Related Work

### Weak supervision

Weak supervision seeks to use domain knowledge and subject matter expertise in opportunistic ways to assign (somewhat noisy) labels to unlabeled data or generate synthetic data. Several general approaches to weak supervision exist. One well-known technique is *distant supervision*^13^, based on using domain knowledge in external knowledge bases. While often used for relation extraction^13, 14^, it has also been used for classification tasks applied to RCT publications^15, 16^. For example, risk of bias judgements in the Cochrane database of systematic reviews were used to automatically label sentences in RCT publications and train models for assessing risk of bias in the publications^15^.

Another increasingly popular weak supervision approach is *data augmentation*. The goal of data augmentation is to increase a model’s generalizability by generating realistic data from a limited number of existing examples. First proposed in computer vision research^17^, it has more recently been adopted in NLP research as well^12, 18^. For example, simple transformations of individual sentences (e.g., synonym replacement, random insertion/deletion) were used to generate additional data and improve modeling accuracy with small datasets^12^. Similar approaches have been adapted to biomedical domain, for tasks ranging from medical abbreviation recognition^19^ to semantic textual similarity^20^ and named entity recognition^21^.

Snorkel has been proposed as a general weak supervision framework^10, 11^. Based on *data programming* paradigm, Snorkel relies on user-defined labeling functions (LFs), which are heuristic methods that can noisily label large quantities of unlabeled data, learns a generative model over the labeling functions to estimate their accuracy and correlations, and generates probabilistic labels that can be used to train machine learning models. Snorkel has been applied to several biomedical text mining tasks, outperforming distant supervision baselines and approaching hand supervision^10^. Other weak supervision approaches have also been developed for biomedical NLP tasks, including smoking status classification from clinical notes^22^, semantic indexing^23^, and clinical entity classification^24^.

### Text mining on RCT publications

Text mining on RCT literature has mostly focused on annotating and extracting study characteristics relevant for systematic reviews and evidence synthesis^25, 26^. PICO elements received much attention; several corpora have been developed at the sentence and span level^16, 27, 28^, and a variety of traditional machine learning methods and deep learning models have been developed to extract these elements from abstracts or full text^16, 28–30^. There is less research on non-PICO elements. Most notably, RobotReviewer^15^ focuses on risk of bias assessment and classifies RCT publications as high or low risk on several risk categories, including sequence generation and allocation concealment. ExaCT^31^ identifies 21 elements in clinical trial publications including sample size and drug dosage. Recently, we constructed a corpus of 50 RCT publications (named CONSORT-TM) annotated at the sentence level with 37 fine-grained CONSORT checklist items to assist with transparency assessment^7^. We also developed baseline NLP models to recognize 17 methodology-specific CONSORT items: two rule-based methods (one keyword-based and another section header-based) as well a linear SVM classifier and a BioBERT-based model. The BioBERT model performed best overall (micro precision: 0.82, recall: 0.63, and F_1_: 0.72), although it failed to recognize infrequent items, which partly motivated this study.

## Materials and Methods

We explored weak supervision to improve the classification performance of our best-performing BioBERT model^7^. In this section, we first describe the collection and pre-processing of unlabeled RCT data from PubMed Central (PMC) for weak supervision. Second, we provide a brief description of the baseline BioBERT models. Third, we discuss our methodology for generating labels using Snorkel framework as well as UMLS-EDA algorithm. Lastly, we provide details on evaluation. The overall procedure is illustrated in Figure 1.

**Figure 1:**
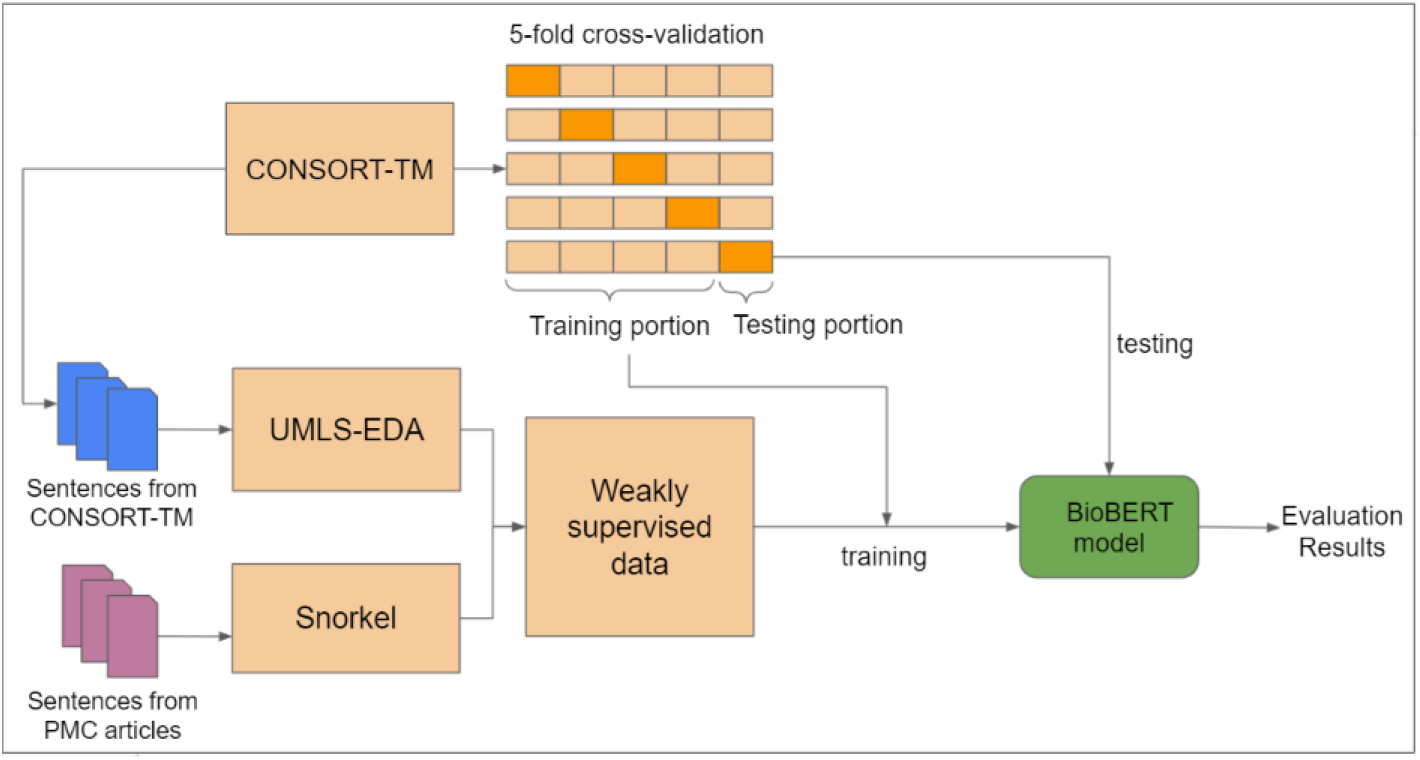
Training and evaluation with weakly supervised data.

### Data collection and pre-processing

We followed the data collection strategy used in previous work^7^ to obtain a large set of RCT articles. Cochrane precision-maximizing search query^b^ was used on 1/15/2021 to search PMC Open Access subset (PMC-OA) for RCT articles published between 1/1/2011 and 12/31/2020^c^. The results were further limited to articles that have full-text XML in PMC-OA. To get a more reliable RCT subset (since publication types in PubMed can be inaccurate), we filtered the results through RCT Tagger^32^, a machine learning model that determines whether a publication is a RCT or not. Its accuracy was found to be 99.7% in predicting RCT studies included in Cochrane systematic reviews^33^. Lastly, we eliminated publications with the word *protocol* in their title (generally study protocol publications).

We used NCBI e-utilities API^d^ to retrieve publications in XML format, and split them into sentences using our in-house sentence splitter^34^. Only sentences that belong to Methods section of the publications were taken into account. Stanford CoreNLP package was used for tokenization and part-of-speech tagging^35^. We eliminated the sentences meeting the following criteria from further consideration, since they are unlikely to indicate CONSORT methodology items: a) contains fewer than five tokens; b) contains numbers only; and c) is a section header or a table/figure caption.

### Baseline models

Our best-performing classifier in previous work^7^ was a BioBERT-based sentence classification model. This classifier uses the BioBERT pretrained language model^8^ as a sentence encoder, considering the model’s output for the [CLS] token as the sentence representation, and trains a sigmoid layer for multi-label classification of 17 CONSORT methodology items. The input to the model is the raw sentence text prepended with its subsection header. The classifier was implemented using `simpletransformers` package^e^. We refer to this model as BASELINE below.

In this study, we used the `huggingface`^f^ BERT implementation. While mostly using the same hyperparameters as BASELINE (batch size: 4, number of epochs: 30, optimizer: Adam, dropout: 0.1), we modified two hyperparameters. First, we used adaptive learning rate instead of a fixed learning rate to optimize the algorithm with different rates based the model performance during training. Second, we set the gradient accumulation steps to 1 (16 for BASELINE), which increases the frequency of model parameter updates. We refer to this optimized model as BASELINE OPT below.

### Generating weak labels using Snorkel

Snorkel^10, 11^ generates weak labels in three steps: a) LF construction; b) creation of a generative model to capture label agreements/disagreements; and c) generation of probabilistic labels for sentences. Input for Snorkel pipeline are unlabeled sentences from RCT publications extracted from PMC-OA.

LFs are expert-defined heuristic rules that can be used to label sentences. For NLP tasks, these can be based on text patterns, syntactic structure, or external knowledge bases. In general, LFs that have high coverage and low overlap are desirable. Such LFs apply to many instances in the dataset yet are unique enough to distinguish instances with different labels. In this study, we used three LF approaches to label CONSORT items: keyword-based, section header-based, and sentence similarity-based. 17 individual LFs were created for each approach (one corresponding to each label).

#### Keyword-based LFs

These LFs mimic the keyword-based method used in previous work^7^. Each CONSORT item is associated with a set of keywords or phrases (e.g., *power to detect* with Sample Size Determination (7a)). A total of 232 phrases are used. Each LF checks whether an input sentence contains one of its keyphrases, and if so, returns the corresponding label as a weak label (or NO-LABEL, if the sentence does not contain a relevant keyword/phrase).

#### Section header-based LFs

These LFs also mimic a baseline method from earlier work^7^. In this case, common subsection headers in Methods sections are associated with CONSORT labels. 48 section header key words/phrases are mapped to CONSORT items (e.g., the word *concealment* to the item Allocation Concealment (9)). These LFs check whether the header of the section to which the sentence belongs matches one of the relevant key phrases.

#### Sentence similarity-based LFs

These LFs assign weak labels to unlabeled sentences based on their similarity to a small set of “ground truth” sentences (a set of 95 sentences provided as examples for specific checklist items in the CONSORT Explanation and Elaboration document^1^ and the CONSORT website^g^). We used BioBERT to generate low-dimensional vector representations of these sentences. For a given unlabeled sentence, we calculate its cosine similarity with every ground truth sentence and consider two labels based on similarity scores: the label of the sentence with the highest similarity and the label that appears most frequently for the top 10 most similar ground truth sentences. If two labels are the same, we use it as the sentence label. Manual checks showed this combination to be more accurate than the most similar sentence label only.

Snorkel applies all LFs to generate a LF matrix that shows the coverage, overlaps, and conflicts between the LFs. *Coverage* information indicates the fraction of the dataset that a particular LF is applied. *Overlap* information shows the fraction of dataset where a particular LF and at least one other LF agree. *Conflict* indicates the fraction of dataset where a particular LB and at least one other LF disagree. Snorkel pools noisy signals from the these three features into a generative model to learn the agreements and disagreements of the LFs, thus assessing the weights of accuracy for each LF. The model then takes into account these accuracies to make a final prediction of the label for each sentence.

### Generating synthetic data using UMLS-EDA

Several CONSORT items are infrequently reported, as they are contingent upon changes in the trial, which may or may not occur (e.g., Changes to Trial Design (3b)^h^). In previous work, text mining methods yielded poor results for these classes^7^, as may be expected. BASELINE model, although it performed best overall in terms of micro-averaging, yielded no predictions for five labels (out of 17) and less than 0.5 F_1_ score for 11 items. We do not expect Snorkel to provide significant number of examples for infrequently reported items, since they are also likely to be rare in the unlabeled dataset and Snorkel’s generative model relies on LF agreement, also likely to be uncommon for such labels.

Therefore, we sought to improve the classification performance for such infrequently reported labels using data augmentation. Specifically, we used UMLS-EDA^21^ and leveraged UMLS synonyms to generate sentences that are similar to the training instances in CONSORT-TM. We define a class as *rare* if the class frequency in the original dataset (*f*) is under a pre-determined threshold (*t*). In generating additional instances, we make up the difference between the frequency in the original dataset and the threshold (i.e., *t* - *f* instances generated) to make the distribution of the training dataset more uniform. If a class is not rare in the original dataset (i.e., *f* >= *t*), no sentences are generated for that label.

UMLS-EDA uses five operations to augment data. *Synonym replacement using WordNet* randomly chooses *n* words from the given sentence that are not stopwords and replaces each with a synonym randomly chosen from WordNet. *Random insertion* inserts random WordNet synonyms of *n* words in the sentence in random positions. *Random swap* randomly swaps the position of two words and repeats this *n* times. *Random deletion* samples and deletes *n* words according to a uniform distribution. *Synonym replacement using UMLS*^*36*^ identifies all the UMLS concepts in the sentence and randomly replaces *n* words in the sentence with a UMLS synonym, also randomly selected. Operations of UMLS-EDA data augmentation are illustrated on an example sentence in Table 1. The parameter *n* is determined dynamically based on the sentence length (*l*) and the operation type (*n* = 0.5\**l* for synonym replacement with UMLS at most and *n* = 0.2\**l* for others). While UMLS-EDA aims to generate *t*-*f* instances, in most cases, a larger number of instances are generated using these operations and we subsample from the generated instances to reach the threshold.

**Table 1:**
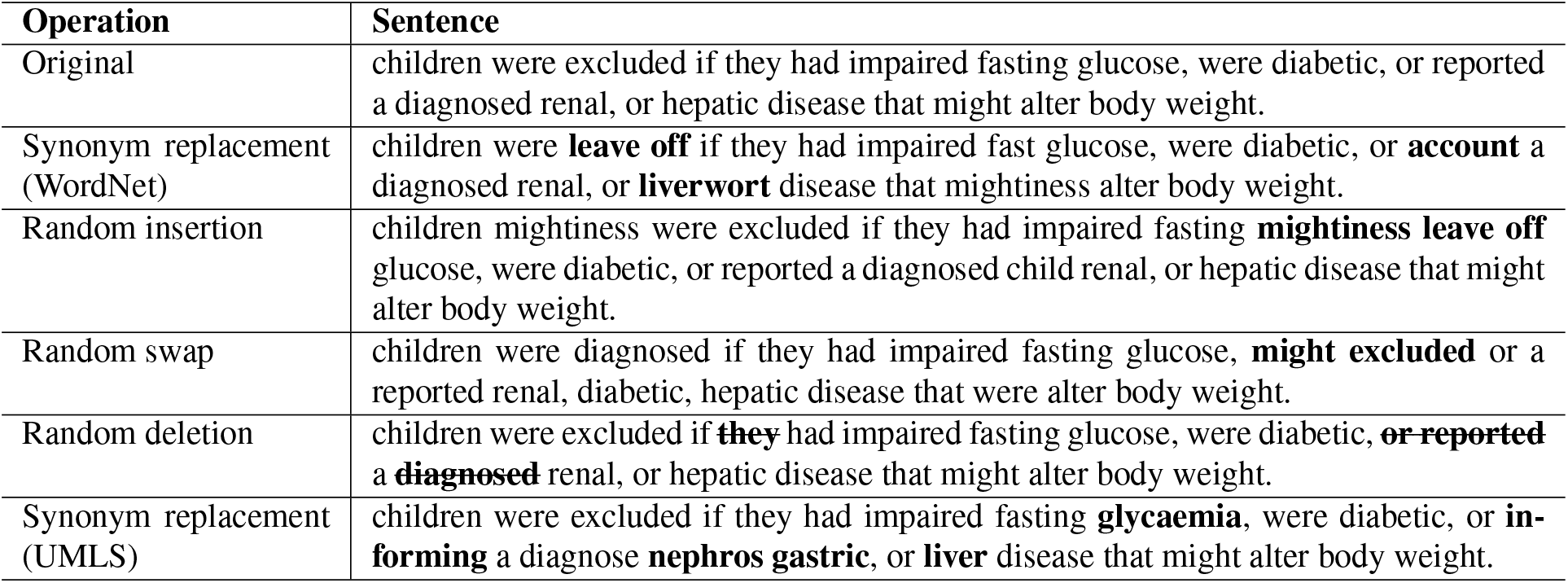
Example of data augmentation using UMLS-EDA. Bold words indicate modifications made by UMLS-EDA. The label of the original sentence is Eligibility Criteria (4a).

### Evaluation

We evaluated whether weak supervision strategies generated labels useful for improving sentence classification performance. For this purpose, we compared the results obtained with BASELINE model on the CONSORT-TM dataset using 5-fold cross validation to results obtained when weakly labeled examples from different strategies are added to the training portion of the folds in cross validation. Note that in this setup, data used for validation and testing in each fold remain the same for all the models. As in previous work, we used standard metrics for evaluation: precision, recall, and their harmonic mean, F_1_ score. In addition to calculating these measures per CONSORT item, we also report micro- and macro-averaged results and the area under ROC curve (AUC).

## Results

### Weak supervision using Snorkel

Our search strategy retrieved a total of 608K RCTs from PubMed, with 155,183 publications having XML full text in PMC. RCT Tagger predicted 71,948 of these as RCTs. Considering only those predicted with a confidence score over 0.95 reduced the dataset to 14,534 publications. Further eliminating publications with *protocol* in the title, we obtained a set of 11,988 papers. A total of 721,948 sentences from these publications was reduced to 551,936 sentences after the filtering approaches discussed above were applied.

We processed 551,936 unlabeled sentences using the Snorkel model, which resulted in 17 probabilities generated for each sentence by the model. We empirically set a probability threshold of 0.8 to predict the final weak labels for the unlabeled sentences. If no label was predicted with a probability higher than 0.8, no label was assigned. The distribution of weak labels generated by Snorkel are shown in Table 2. Noting that most weak labels corresponded to items that are already relatively well-represented in the dataset, we limited the number of weakly labeled examples for each CONSORT item to a pre-determined threshold in our classification experiments and randomly sampled these examples. We report the results with the threshold that performed best in our experiments (500).

**Table 2:**
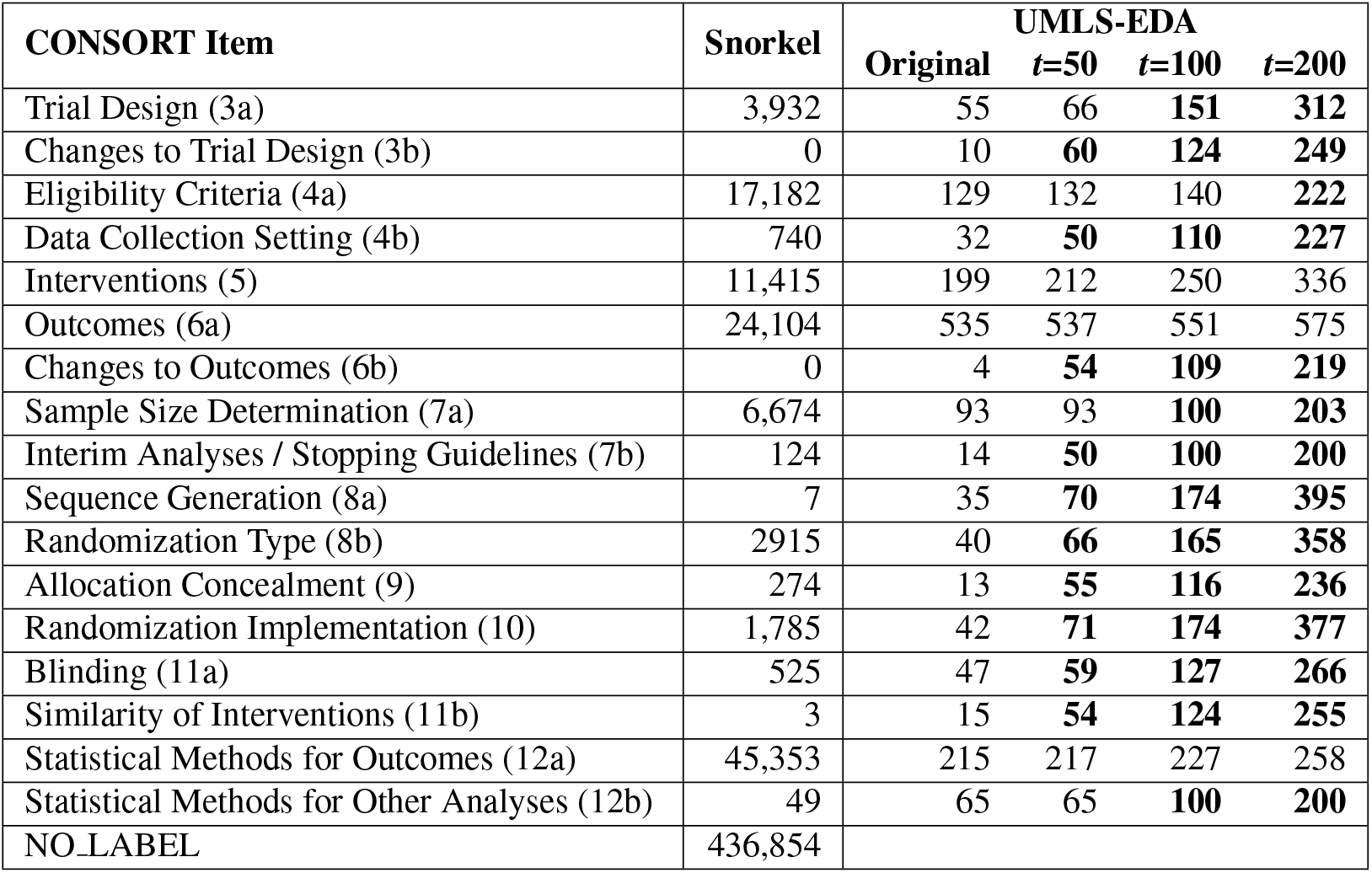
The frequency of each methodology item in CONSORT-TM and the augmented data generated by Snorkel and UMLS-EDA. Highlighted numbers correspond to the cases when the CONSORT item was considered a rare label and augmented for the given threshold *t*.

| CONSORT Item | Snorkel | UMLS-EDA |  |  |  |
| --- | --- | --- | --- | --- | --- |
| | | Original | $t=50$ | $t=100$ | $t=200$ |
| Trial Design (3a) | 3,932 | 55 | 66 | **151** | **312** |
| Changes to Trial Design (3b) | 0 | 10 | **60** | **124** | **249** |
| Eligibility Criteria (4a) | 17,182 | 129 | 132 | 140 | **222** |
| Data Collection Setting (4b) | 740 | 32 | **50** | **110** | **227** |
| Interventions (5) | 11,415 | 199 | 212 | 250 | 336 |
| Outcomes (6a) | 24,104 | 535 | 537 | 551 | 575 |
| Changes to Outcomes (6b) | 0 | 4 | **54** | **109** | **219** |
| Sample Size Determination (7a) | 6,674 | 93 | 93 | **100** | **203** |
| Interim Analyses / Stopping Guidelines (7b) | 124 | 14 | **50** | **100** | **200** |
| Sequence Generation (8a) | 7 | 35 | **70** | **174** | **395** |
| Randomization Type (8b) | 2915 | 40 | **66** | **165** | **358** |
| Allocation Concealment (9) | 274 | 13 | **55** | **116** | **236** |
| Randomization Implementation (10) | 1,785 | 42 | **71** | **174** | **377** |
| Blinding (11a) | 525 | 47 | **59** | **127** | **266** |
| Similarity of Interventions (11b) | 3 | 15 | **54** | **124** | **255** |
| Statistical Methods for Outcomes (12a) | 45,353 | 215 | 217 | 227 | 258 |
| Statistical Methods for Other Analyses (12b) | 49 | 65 | 65 | **100** | **200** |
| NO_LABEL | 436,854 |  |  |  |  |

### Weak supervision using UMLS-EDA

We used thresholds 50, 100, and 200 to generate on average 246, 844, and 2217 additional examples, respectively, using UMLS-EDA. Data augmentation was implemented as part of 5-fold cross-validation, and therefore, number of examples between folds differ. The numbers of instances for each label in the original dataset and the augmented datasets (for one of the folds) are shown in Table 2. Note that a label can be considered *rare* or not at different threshold values and may or may not be augmented. For example, while the item Statistical Methods for Other Analyses (12b) is not rare when the threshold is 50, it is considered rare for the threshold 100 and, therefore, augmented (Table 2). The number of rare class instances generally do not add up to the threshold exactly, because it is possible to label an augmented example with more than one class.

### Classification results

We trained and tested BASELINE and BASELINE OPT models on CONSORT-TM using 5-fold cross-validation. In other experiments, we used various sizes of weakly supervised data obtained using Snorkel and UMLS-EDA as additional training data. For brevity, we only report the results for the best-performing model-data size combination. For Snorkel, this is BASELINE OPT model augmented with maximum 500 examples per label. For UMLS-EDA, it is the same model augmented with UMLS-EDA data with a threshold of 50. The results (Table 3) show that hyperparameter tuning (BASELINE OPT) makes a significant difference in performance (7% increase in micro-F_1_ and 63% increase in macro-F_1_), while the impact of weak supervision strategies seems minor; Snorkel data leads to a slight performance degradation, while UMLS-EDA data increases micro-F_1_ by one percentage point and AUC with 1.6 points, with practically no change in macro-F_1_.

**Table 3:** Classification results using CONSORT-TM and weakly supervised data. SNORKEL(500) uses BASE-LINE OPT with additional 500 instances per label from Snorkel data. UMLS-EDA(50) uses BASELINE OPT with additional instances from UMLS-EDA to add up to at least 50 instances for each label. 3a: Trial Design; 3b: Changes to Trial Design; 4a: Eligibility Criteria; 4b: Data Collection Setting; 5: Interventions; 6a: Outcomes; 6b: Changes to Outcomes; 7a: Sample Size Determination; 7b: Interim Analyses/Stopping Guidelines; 8a: Sequence Generation; 8b: Randomization Type; 9: Allocation Concealment; 10: Randomization Implementation; 11a: Blinding Procedure; 11b: Similarity of Interventions; 12a: Statistical Methods for Outcomes; 12b: Statistical Methods for Other Analyses. P: precision; R: recall; F: F_1_ score; AUC: Area Under Receiver Operator Characteristic (ROC) Curve.

| CONSORT Item | BASELINE |  |  | BASELINE_OPT |  |  | SNORKEL(500) |  |  | UMLS-EDA(50) |  |  |
| --- | --- | --- | --- | --- | --- | --- | --- | --- | --- | --- | --- | --- |
|  | P | R | F | P | R | F | P | R | F | P | R | F |
| 3a | 0.93 | 0.49 | 0.63 | 0.83 | 0.82 | 0.82 | 0.74 | 0.77 | 0.75 | 0.82 | 0.75 | 0.78 |
| 3b | 0.00 | 0.00 | 0.00 | 0.00 | 0.00 | 0.00 | 0.00 | 0.00 | 0.00 | 0.00 | 0.00 | 0.00 |
| 4a | 0.90 | 0.82 | 0.85 | 0.91 | 0.87 | 0.89 | 0.88 | 0.88 | 0.88 | 0.93 | 0.88 | 0.90 |
| 4b | 0.80 | 0.24 | 0.36 | 0.93 | 0.84 | 0.87 | 0.81 | 0.80 | 0.79 | 0.83 | 0.80 | 0.81 |
| 5 | 0.76 | 0.69 | 0.72 | 0.76 | 0.75 | 0.75 | 0.73 | 0.76 | 0.73 | 0.76 | 0.76 | 0.75 |
| 6a | 0.84 | 0.78 | 0.81 | 0.81 | 0.83 | 0.82 | 0.83 | 0.83 | 0.83 | 0.83 | 0.83 | 0.83 |
| 6b | 0.00 | 0.00 | 0.00 | 0.00 | 0.00 | 0.00 | 0.00 | 0.00 | 0.00 | 0.00 | 0.00 | 0.00 |
| 7a | 0.88 | 0.80 | 0.84 | 0.90 | 0.88 | 0.88 | 0.87 | 0.93 | 0.90 | 0.90 | 0.90 | 0.90 |
| 7b | 0.00 | 0.00 | 0.0 | 0.92 | 0.61 | 0.70 | 0.72 | 0.68 | 0.70 | 0.95 | 0.71 | 0.78 |
| 8a | 0.86 | 0.26 | 0.38 | 0.92 | 0.87 | 0.88 | 0.86 | 0.69 | 0.76 | 0.91 | 0.87 | 0.88 |
| 8b | 0.71 | 0.29 | 0.38 | 0.75 | 0.72 | 0.73 | 0.65 | 0.71 | 0.67 | 0.81 | 0.70 | 0.75 |
| 9 | 0.00 | 0.00 | 0.00 | 0.58 | 0.46 | 0.45 | 0.41 | 0.41 | 0.40 | 0.54 | 0.51 | 0.43 |
| 10 | 0.72 | 0.15 | 0.24 | 0.61 | 0.48 | 0.53 | 0.48 | 0.56 | 0.50 | 0.55 | 0.50 | 0.52 |
| 11a | 0.77 | 0.29 | 0.42 | 0.71 | 0.65 | 0.66 | 0.60 | 0.61 | 0.59 | 0.744 | 0.61 | 0.66 |
| 11b | 0.00 | 0.00 | 0.00 | 0.60 | 0.44 | 0.45 | 0.70 | 0.31 | 0.41 | 0.57 | 0.44 | 0.44 |
| 12a | 0.75 | 0.76 | 0.75 | 0.75 | 0.81 | 0.77 | 0.74 | 0.84 | 0.78 | 0.74 | 0.83 | 0.78 |
| 12b | 0.05 | 0.03 | 0.04 | 0.40 | 0.30 | 0.32 | 0.40 | 0.21 | 0.24 | 0.48 | 0.26 | 0.31 |
| Micro-average | 0.82 | 0.63 | 0.72 | 0.79 | 0.76 | 0.77 | 0.76 | 0.75 | 0.76 | 0.80 | 0.76 | 0.78 |
| Macro-average | 0.52 | 0.33 | 0.38 | 0.67 | 0.61 | 0.62 | 0.61 | 0.58 | 0.58 | 0.67 | 0.61 | 0.62 |
| AUC |  | 0.812 |  |  | 0.876 |  |  | 0.875 |  |  | 0.892 |  |

## Discussion

### Weak supervision with Snorkel

Approximately 21% of unlabeled sentences were assigned a weak label by Snorkel. The number of weakly assigned labels reflected to some extent the distribution of labels in the original dataset; many sentences were weakly labeled with common labels, such as Outcomes (6a). On the other hand, Snorkel failed to weakly label any sentences with two most infrequent labels (Table 2).

The quality of Snorkel labels depends largely on the quality of LFs. We used two LFs based on heuristics explored in previous work. Micro-F_1_ for both methods were found to be around 0.50 in previous work (0.50 for keyword-based and 0.45 for section header-based). More accurate LFs could lead to better models of LF behavior, improving Snorkel results. To better understand the quality of Snorkel-generated weak labels, we sampled 318 sentences and two authors of this paper (LH and HK) independently labeled the sentences, without access to Snorkel labels. We calculated the agreement of these annotations with Snorkel-generated labels, using Krippendorff’s *α* with the distance metric MASI^37^ which accounts for partial agreement in the case of multiple labels. *α* agreements between Snorkel and each annotator were found to be 0.46 and 0.61, respectively. Inter-annotator agreement was 0.59. Interestingly, agreement between Snorkel and simple majority vote was 0.93. Agreement results suggest that Snorkel may converge to this simple heuristic in some cases, and that it behaves more or less like another annotator in the process.

We found that a large percentage of annotator disagreement with Snorkel came from randomization-related labels (items 8a, 8b, 9, and 10). These items often appear in the same sentence and the clues for them can be overlapping, making it a challenge to label them accurately for both humans and automated methods. In previous work, we found inter-annotator agreement for these items to be somewhat low as well (*α*=0.62, 0.48, 0.34, 0.35, respectively)^7^. Snorkel tends to pick a single label for sentences, and this was especially problematic for randomization-related sentences.

### Weak supervision using UMLS-EDA

Data augmentation is expected to reduce overfitting and help with model robustness^17^. While generating data using UMLS-EDA is relatively cheap, the resulting sentences are generally not meaningful, making it difficult to assess the quality of the augmented data (in contrast to Snorkel), aside from the downstream model performance that it produces. We make several observations based on our examination of the augmented data. One of the data augmentation operations (synonym replacement with UMLS) may need to be refined. UMLS synonyms that are used to replace the original words/phrases are sometimes different from the original only in trivial ways (acronyms or swapped tokens), and strategies that only allow more significant replacements could be beneficial. For example, it might be worthwhile to limit the replacement only to terms of particular semantic types or part-of-speech tags. Similar observations were made for synonym replacement with WordNet, as well. Some replacements involved functional words, which may not be as beneficial as replacing content words (nouns, adjectives).

### Effect of weak supervision and model hyperparameters on classification performance

We did not observe significant improvements in classification performance due to weakly supervised data. Neither strategy led to any correct predictions for the two most infrequent labels (3a, 6a). While this was not unexpected in the case of Snorkel (as no additional examples were labeled with these items), it was more surprising in the case of UMLS-EDA, which seemed to generate sufficient number of examples for these items. We observed AUC improvement with UMLS-EDA (0.892 vs. 0.876 with BASELINE OPT), which may indicate that UMLS-EDA helps with robustness and generalizability. As UMLS-EDA approach is cheap, further refinements to it may be promising as a future direction.

Somewhat to our surprise, we found that model hyperparameters made a much more significant difference in model performance. BASELINE OPT model yielded about 7% improvement in micro-F_1_ and 63% improvement in macro-F_1_ over the BASELINE model, with improvements in almost all labels. To assess how hyperparameters interacted with weak supervision, we also measured performance when BASELINE model (instead of BASELINE OPT) was trained with weakly supervised data. Using Snorkel for weak supervision in this scenario improved micro-F_1_ from 0.72 to 0.75, suggesting that hyperparameter optimization may, in some cases, obviate the need for additional (noisy) data.

### Limitations

Our investigation was limited to one relatively small corpus. The findings regarding weak supervision (as well as Snorkel and UMLS-EDA specifically) may not be generalizable to other corpora. We used few heuristics with modest performance as LFs and Snorkel label quality is likely to be improved with with additional more accurate LFs; however, this requires some domain expertise. While we performed some hyperparameter tuning, we did not do an exhaustive search for optimal parameters, and it is possible that more optimal hyperparameters can improve results further.

## Conclusions and future work

We investigated the impact of two weak supervision strategies on multi-label sentence classification models of RCT publications. We did not observe a clear positive impact of weak supervision on the specific task we studied. More experiments would be needed to determine whether this is a corpus-specific finding or it is more general. Various forms of weak supervision has been shown to improve classification performance^12, 16^, mostly in multi-class cases; therefore, it is possible that our weak supervision strategies need more refinement for the multi-label case.

In future work, we plan to refine our approach. For example, in UMLS-EDA, we can devise methods to generate more contextually appropriate synonyms from WordNet and UMLS. Snorkel would benefit from more accurate LFs. Other semi-supervised learning approaches (e.g., self-training^38^, few-shot learning^39^) can also be investigated as alternatives.

## Data Availability

An annotated corpus used in this work,CONSORT-TM, is publicly available at https://github.com/kilicogluh/CONSORT-TM.

## Footnotes

b https://work.cochrane.org/pubmed

c The start date is chosen based on the most recent publication of CONSORT guidelines (2010)^1^.

d https://https://www.ncbi.nlm.nih.gov/books/NBK25501/

e https://github.com/ThilinaRajapakse/simpletransformers

f https://huggingface.co/

g http://www.consort-statement.org/examples/sample

h We use the item numbers used in CONSORT guidelines, as well, hereafter.

